# Australian medical research that considered sex as a biological variable: a meta-analysis

**DOI:** 10.1101/2024.08.23.24310791

**Authors:** Janelle Ryan, Shanie Landen, Vincent Harley

## Abstract

Subjects in medical research have predominantly been male (1). Women experience 50-75% more adverse drug responses (2) resulting in withdrawn medications (3). While sex differences in metabolism, disease and treatment response are increasingly recognised, sex-informed medicine is lagging. In 2016, USAs National Institutes of Health (NIH) formulated the Sex as a Biological Variable policy (4), stating that grant recipients must consider sex in experimental design, planning, analysis and reporting of their findings. Australian data is lacking on the inclusion of both males and females as well as appropriate analysis of sex differences.

We analysed the 219 Medical Journal of Australia (MJA) research articles over 2019-2023 (Box 1). We tallied when; i) both males and females were included in the study, ii) sex differences were reported and/or considered, and iii) the analysis was appropriate to support sex-related claims.

We found that articles published in MJA are including males and females, however testing of sex differences is uncommon and appropriate statistical analysis is lacking. We hope that this article will bring attention to this fundamental issue and improve future efforts to investigate sex differences.

## Research Letter

Subjects in medical research have predominantly been male (1). Women experience 50-75% more adverse drug responses (2) resulting in withdrawn medications (3). While sex differences in metabolism, disease and treatment response are increasingly recognised, sex-informed medicine is lagging. Only one medication has sex-specific prescription (NDA021774). In 2016, USA’s National Institutes of Health (NIH) formulated the “Sex as a Biological Variable” policy (4), stating that grant recipients must consider sex in experimental design, planning, analysis and reporting of their findings. Australian data is lacking on the inclusion of both males and females as well as appropriate analysis of sex differences. Here, ‘sex” refers to biological attributes associated typically with a combination of physical features (i.e. male and female). “Gender” refers to socially constructed roles and identities (i.e. man and woman).

We analysed the 219 *Medical Journal of Australia (MJA)* research articles over 2019-2023 (Box 1). We tallied when; i) both males and females were included in the study, ii) sex differences were reported and/or considered, and iii) the analysis was appropriate to support sex-related claims.

**Box 1.**
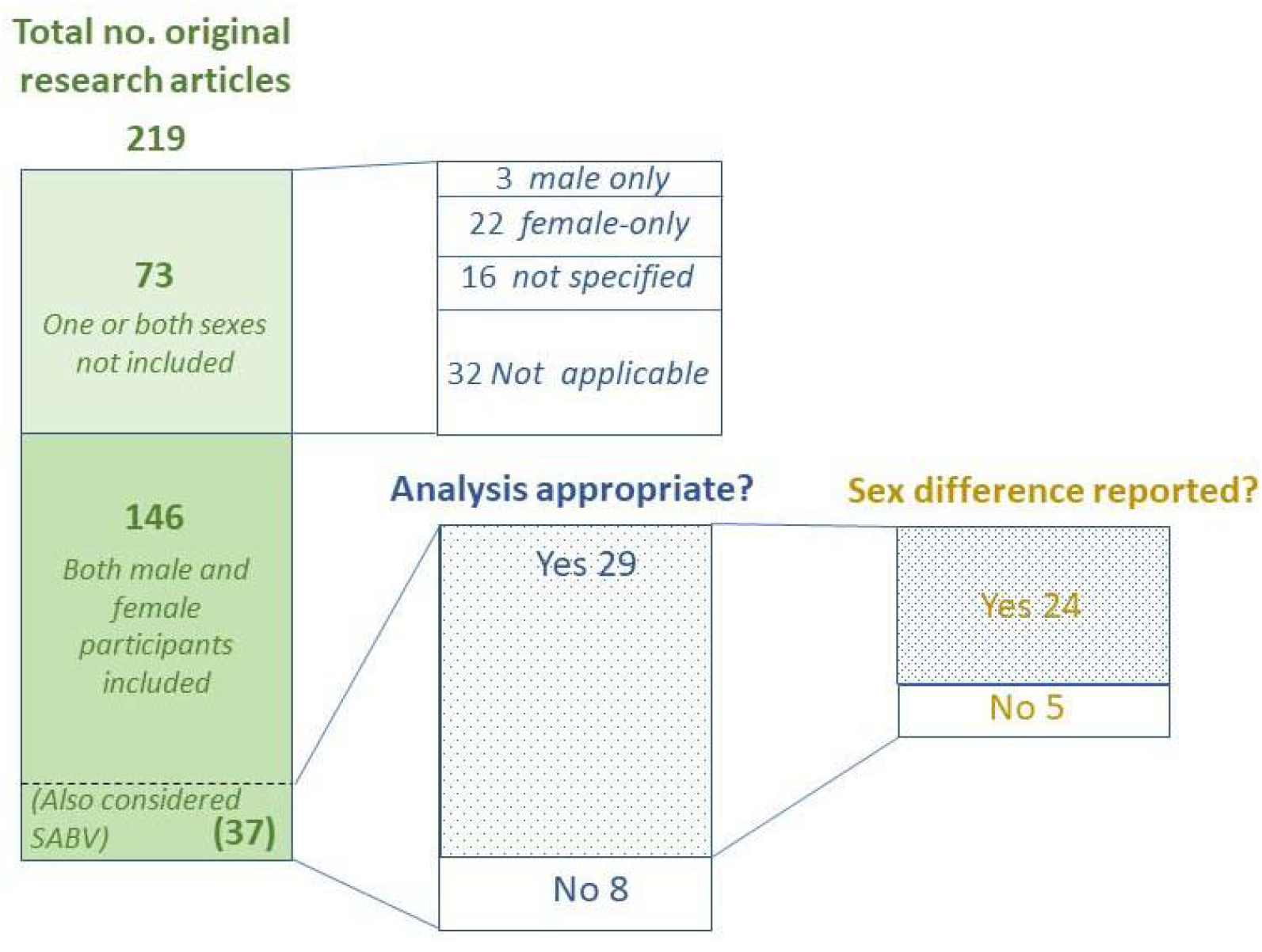
Analysis of Original Research Articles in the Medical Journal of Australia in the last five years (2019-2023). Of 219 Original Research Articles, 146 included both male and female participants. 37 of those also considered sex as a biological variable.

146 of 219 articles (67%) included both male and female participants (Box 1). The remaining 73 (33%) were appropriately single-sex disease studies (ie. ovarian cancer), or studies where sex was not applicable (e.g. computer modelling) (Box 1). Over the five-year period, there has been little change in this proportion including both males and females [29/44 (65%) in 2019 versus 31/46 (67%) in 2023 (Appendix, Table 1)].

37 of 146 articles (25%) tested for sex differences (Box 1). 29 of 37 articles (78%) used statistics appropriately by considering sex as a covariate, and/or testing for an interaction between sex and treatment/intervention variables (Box 1). 24 of 29 articles (83%) employing appropriate tests reported a sex difference. Of concern is the 7/37 (19%) of *MJA* articles that reported a sex difference but did not employ appropriate statistical analysis to support sex difference claims.

In a meta-analysis of 34 high impact international journals, the proportion of studies including both males and females increased from 28% in 2009 (841 studies) to 49% in 2019 (720 studies) (5). *MJA*’s ∼65% compares favourably against this ∼49% international benchmark. In the same study, the proportion of studies testing for sex differences remained steady at ∼50% from 2009 to 2019 (5), thus the fraction of articles testing for sex differences in Australia (25%) is about half that of the international benchmark. One explanation may be that some studies are underpowered to consider sex differences. A different study of selected US journals found only 27/92 (29%) of articles that reported a sex difference in 2019 employed appropriate statistical analysis (appropriate statistical methods outlined in Box 2) (6). By comparison, we found 24/29 (83%) employed an appropriate statistical test.

**Box 2.**
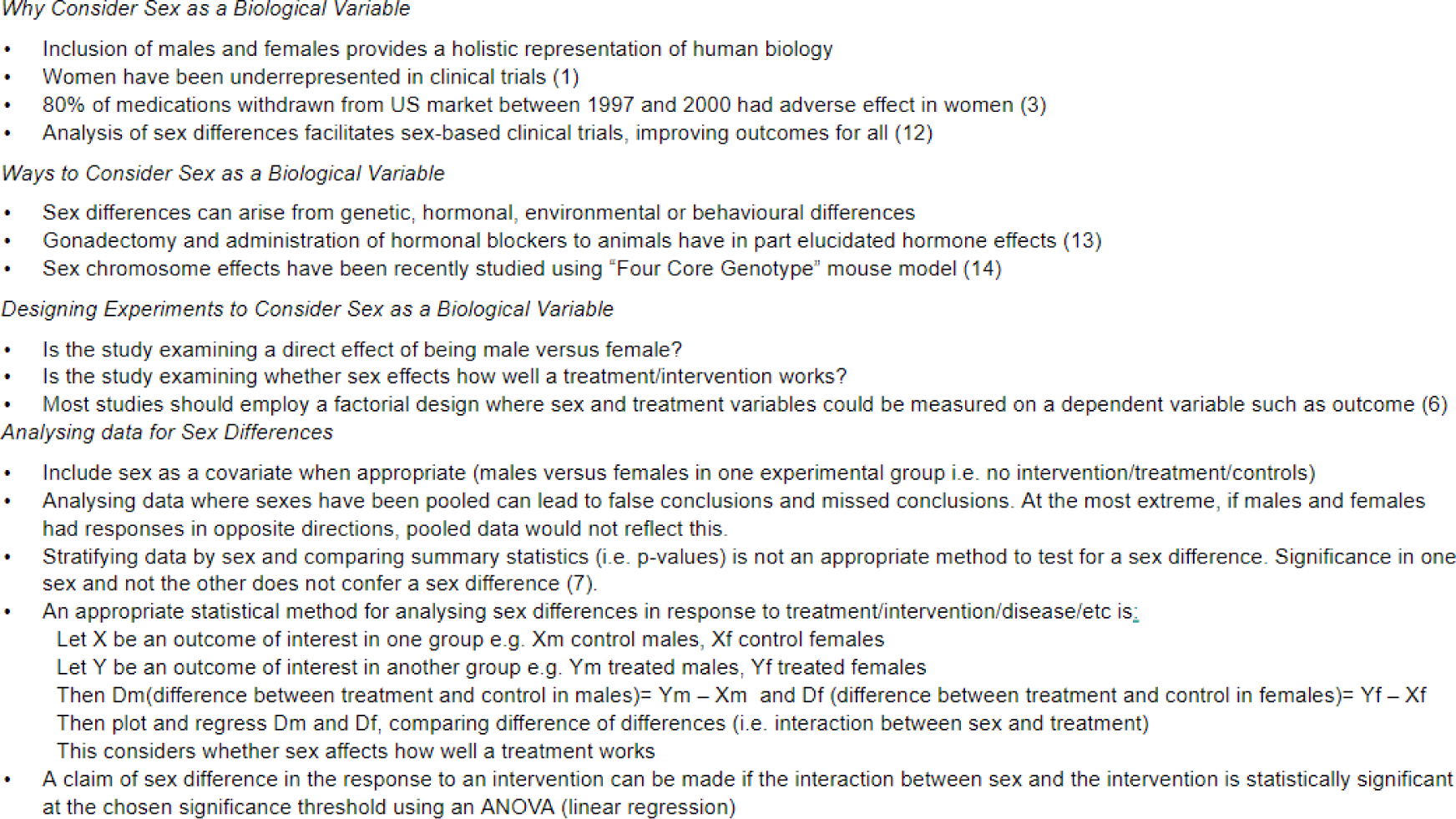
Recommendations for considering sex as a biological variable in experimental design and analysis.

We noted that gender-related language (men and women) was often used interchangeably with sex related-language (males and females). This conflation can make assessing a claim of sex difference difficult given a person’s gender identity and sex at birth may be incongruous. We recommend that authors be specific about how they define sex and/or gender.

Researchers often stratify each sex during analysis when comparing the effect of a given intervention/treatment and claim that significance in one sex but not the other qualifies as a sex difference. This was a common error we observed in our analysis (6/7 articles, 86%). Without making a direct statistical comparison, sex differences cannot be analysed and this mistake has been reviewed elsewhere (6,7). Appropriate statistical methods when testing for sex differences are outlined in Box 2.

Compared with global benchmarks, articles published in *MJA* are including both males and females in research, but analysis of sex differences remains uncommon, with appropriate analysis also lacking. This suggests a need for a national policy on inclusion of sex as a biological variable. While Australian funding agencies are yet to address sex in research policy (8), efforts are ongoing, with the National Health and Medical Research Council/Medical Research Future Fund releasing a draft policy for public consultation in 2023. The funding agencies are pivotal in driving policy change. Australia should ensure that sufficient funding is provided for research and clinical trials to be powered appropriately. However, the US experience has shown that policy alone will not increase the integration of sex and gender related analysis into health and medical research (6). There is a need for specialised training and public health and medical curricula on study design, analysing and interpreting research that considers sex as a biological variable. In lieu of formal guidance, published roadmaps can assist (9). Beyond inclusion of sex and gender in research and descriptions of any differences, researchers should consider intersectionality-based analysis and establishing the origin (10,11).

In summary, we analysed whether *MJA* research articles published over the last five years considered sex as a biological variable. We found that articles published in *MJA* are including males and females, however testing of sex differences is uncommon and appropriate statistical analysis is lacking. We hope that this article will bring attention to this fundamental issue and improve future efforts to investigate sex differences.

## Data Availability

All data produced in the present study are available upon reasonable request to the authors.

## Ethics Approval Statement

Ethics was not sought for this meta-analysis. Only published articles were considered.

## Data Sharing Statement

The study data can be accessed by contacting the corresponding author.

## Funding Statement

This research received financial support from the National Health and Medical Research Council Program Grant 2002426 and Fellowship APP1154870 awarded to VH. Additional support was provided by the Australian Government Research Training Program Scholarship for JR. We also acknowledge funding through the Victorian Governments Operational Infrastructure Support Program.

## Notes

### Competing Interest Statement

The authors have declared no competing interest.

### Author Declarations

We used only published, openly available research articles that reported human data from the Medical Journal of Australia journal.

