## Appendices for "Australian medical research that considered sex as a biological variable: a meta-analysis"

Appendices, Table 1. Articles that included male and female participants by year.

| Year | Number of Articles | Number that included both males and females | Percentage |
| --- | --- | --- | --- |
| 2019 | 44 | 29 | 65.9% |
| 2020 | 44 | 24 | 54.5% |
| 2021 | 41 | 30 | 73.1% |
| 2022 | 44 | 32 | 72.7% |
| 2023 | 46 | 31 | 67.3% |
|  | 219 total | 146 total | 66.7% total |
